# County and Demographic Differences in Drug Arrests and Controlled Substance Use in Maine

**DOI:** 10.1101/2020.08.19.20178079

**Authors:** Trajan F. Hyde, Amadea D. Bekoe-Tabiri, Amalie K. Kropp Lopez, Luis G. Devia, Belsy D. Gutierrez, Matthew C. Lara, Anthony R. Soto, Daniel E. Kaufman, Kevin J. Simpson, Matthew T. Moran, Dipam T. Shah, Michelle Foster, Clare E. Desrosiers, John Herbert, Stephanie D. Nichols, Kenneth L. McCall, Brian J. Piper

**Author notes:** designated authors contributed equally (Trajan F. Hyde, Amadea D. Bekoe-Tabiri). Department of Medical Education, Geisinger Commonwealth School of Medicine, Scranton, PA, or.

## Abstract

**Introduction:** The Diversion Alert Program (DAP) was established to curb the misuse of drugs and help identify people in need of substance use disorder (SUD) treatment. Law enforcement compiled arrest data into a database accessible by health care providers. The objectives of this investigation were to identify regional and demographic differences in drug use and misuse in Maine.

**Methods:** All arrests (N = 11,234) reported to the DAP from 2013 to 2018 were examined by county, arrestee demographics, and classified into families (opioids, stimulants, sedatives). The Automation of Reports and Consolidated Orders System (ARCOS) tracks the distribution of controlled pharmaceuticals (schedule I-V). Opioids were converted to oral morphine mg equivalents (MME). County and zip-code heat maps were constructed.

**Results:** The counties with the most arrests per capita were Cumberland, Androscoggin, and Penobscot. Opioids were the most common drug class in arrests in all counties except Aroostook, where stimulants were most common. Medical distribution of opioids varied. With the exception of buprenorphine, which doubled, many prescription opioids like hydrocodone, fentanyl, and oxymorphone exhibited large (>50%) reductions. Methadone was the predominant opioid statewide (56.4% of the total MME) although there were sizeable differences (Presque Isle = 8.6%, Bangor = 78.9%) and this is likely impacted by use for SUD treatment. Amphetamine distribution increased by 67.9%. *Conclusions:* The DAP was useful to prevent information silos and enhance communication between law enforcement and health care providers. Maine’s DAP was a unique system to identify patients in need of additional treatment resources. The increase in prescription stimulants may warrant continued monitoring.

---

Maine has been particularly impacted by the national opioid crisis and more than 80% of people who use heroin started with prescription opioids.^1^ Opioid prescribing changes over the past two-decades differed appreciably in the US. In states such as West Virginia, Kentucky, and most importantly, Maine, prescribing rates were 2.5 to 5.0 times higher than the national average for hydrocodone and oxycodone.^2^ The prescription morphine milligram equivalent (MME) per person in the New England states were among the highest in the United States. Maine was ranked fifth nationally in 2016 at 1,393 MME, which increased to 2,000 MME when Opioid Use Disorder (OUD) medications, buprenorphine and methadone, were included. States with a higher median age, such as Maine, also used more prescription opioids.^3^ Cumberland county, containing Maine’s largest city of Portland, accounted for 60% more drug related deaths from 1997 – 2002 than would be anticipated based on population indicating an ongoing issue that has been plaguing the region. Methadone, oxycodone, propoxyphene, hydrocodone and other prescription opioids were common drugs mentioned in the death certificates the last two decades.^4^ Drug fatalities in as recent as 2019 continued to involve opioids in 84% of the cases, and typically with other substances including fentanyl analogues which have been on the rise since 2013.^5,6^ Innovative strategies are necessary to address this ongoing public health crisis.

Maine had a novel harm-reduction database, the Diversion Alert Program (DAP) which originated in Aroostook County and was expanded statewide to improve communication between law enforcement and healthcare providers. The DAP included the patient’s name, date of birth, town of residence, drug charge, implicated drugs, and arresting agency for adults whose arrest involved illicit substances, prescription medications and non-prescription pharmaceuticals.^7^ As a point of care tool, this resource could be used to help identify patients that might have needed specialist (e.g. pain, psychiatry, or addiction medicine) involvement. The DAP was also employed as pharmacoepidemiological source with regular reports for 2014 - 2017.^3,5,7,8,9^ The top substances involved in arrests were heroin, cocaine and crack cocaine, buprenorphine, oxycodone, methamphetamine, alprazolam, clonazepam, marijuana, hydocodone, fentanyl, amphetamine, alpha-PVP/bath salts, and gabapentin. Possession accounted for three-fifths of charges followed by trafficking. Older adults (> 60) had a significant and disproportionate percent of their arrests involving oxycodone and hydrocodone.^8^ The annual reports showed that Cumberland and Androscoggin counties led the state on population corrected arrests.^7,8^

The first objective of this study was to provide an analysis of nonmedical use of controlled substance as reported by DAP at a county level. The second objective included examining whether the arrest profile differed by arrestee demographic, specifically their biological sex and age. The third objective was to use a comprehensive data source, the Drug Enforcement Administration’s (DEA) Automation of Reports and Consolidated Ordering System (ARCOS) to determine regional changes in medical use of controlled substances in Maine.

## METHODS

### Procedures

Two complementary data sources were used: the DAP and ARCOS. The sample included all arrests (N = 11,234) reported to the DAP from when the program began (mid-2013) until it ceased operation due to lack of funding (mid-2018). Local, state and federal law enforcement agencies provided information. A de-identified spreadsheet containing information of age, sex, county of arrest, substance, and offense was obtained. Arrest data was classified by the arrestee’s county of residence. State and county totals for the following categories were calculated: total number of arrests, total population, percentage of county population arrested, percent of arrests involving females, mean age of total arrests, percent of arrests involving opioids or stimulants, most frequent federal drug schedule, and most common level (federal, state, county, or city) of arresting agency. For simplicity, county and city agencies are henceforth designated as “local”. Additional information including the processing steps for arrests involving multiple drugs is available elsewhere.^7^

The DEA’s ARCOS is a federal program created with the 1970 Controlled Substances Act that collects data (weight distributed in grams) of Schedule II and III substances distributed to pharmacies, hospitals, methadone treatment programs (referred to by the DEA as narcotic treatment programs) and providers.^10^ Thirteen opioid pain medications were examined from 2008 to 2017: oxycodone, fentanyl, morphine, hydrocodone, hydromorphone, oxymorphone, tapentadol, codeine, meperidine, dihydrocodeine, sufentanil, remifentanil, alfentanil, and two, primarily OUD opioids: methadone and buprenorphine. The oral MME conversion factors (e.g. tapentadol = 0.4, methadone = 10) are available elsewhere.^3^ The weights distributed of stimulants: methylphenidate, amphetamines, lisdexamfetamine specifically, and the barbiturates: pentobarbital and secobarbital were also obtained. ARCOS was previously validated by comparing results for a single opioid with that obtained for the Maine Prescription Monitoring Program which revealed a high correlation.^3^ Procedures were approved by the ethics committee of the University of New England.

### Data-analysis

The percentage of the county population arrests was determined by dividing the total number of county arrests by the county population in 2015 (i.e. the midpoint year).^11^ Maine’s population increased by 1.3% from 2008 to 2017. The percent of female arrests was calculated by dividing the county female arrest total by the total county population. The mean age of total arrests was determined by averaging the ages of the arrests in each county. The percent of arrests involving opioids and stimulants respectively were calculated by dividing the arrests involving each drug class by the total amount of arrests in the county. ARCOS also reports drug distribution by the first three-digits of the zip code. For simplicity, 039 = York, 040 = Kennebunk, 041 = Portland, 042 = Lewiston, 043 = Augusta, 044 = Bangor, 045 = Boothbay Harbor, 046 = Bar Harbor, 047 = Presque Isle, 048 = Rockport, and 049 = Waterville. Relative to their peak year, a change of 0 to 19.9%, 20.0 to 49.9%, and > 50.0% in controlled substance weight per year were interpreted as small, medium, and large respectively. County heat maps were generated with Microsoft Excel and QGIS.

## RESULTS

There were 11,234 drug-related arrests reported to the DAP between 2013 and 2018. The mean age of individuals arrested was highest at 36.4 years old in Somerset compared to Piscataquis at 31.0 years old (Table 1). Androscoggin has a population 107,233 with a total of 1,372 drug arrests making it the county with the highest percentage of the population for drug arrests (1.28%, Table 1; Figure 1A, B). The average drug arrest percentage for the state was 0.78% (Figure 1B). In Androscoggin, 35.2% of the arrested population was comprised of females, compared to Lincoln County where 42.5% of the arrested population was female (Figure 1C). Figure 1D shows the average age of females arrested. Fifteen of the sixteen counties reported more opioid than stimulant arrests (Figure 1E, F). Counties with the greatest percentage of arrests involving opioids were Knox (57.0%), Waldo (56.8%), Kennebec (55.9%) and Hancock (55.6%) (Table 1; Figure 1E). Stimulants accounted for fewer than one-eighth of arrests (11.7%) in Waldo and Lincoln counties versus almost two-fifths (38.7%) in Aroostook (Figure 1F). Schedule II substances (e.g. cocaine) were most common in thirteen (81.25%) counties. State agencies were the agency responsible for the most arrests in over-half (56.3%) of counties (Table 1).

**Table 1.** Maine Diversion Alert Program arrests from 2013-2018.

| **County** | **Number**<br>**of Total**<br>**Arrests** | **Total**<br>**Population**<br>**Size (2015)** | **% of Pop**<br>**w. Drug**<br>**Arrests** | **% of**<br>**Females** | **Age** | **% Total**<br>**Arrests**<br>**Involving**<br>**Opioids** | **% Total**<br>**Arrests**<br>**Involving**<br>**Stimulants** | **Most**<br>**Common**<br>**Federal**<br>**Class** | **Most**<br>**Common**<br>**Arresting**<br>**Agency** |
| --- | --- | --- | --- | --- | --- | --- | --- | --- | --- |
| Androscoggin | 1,372 | 107,233 | 1.28% | 35.2 | 32.4 | 42.1 | 35.9 | 2 | Local |
| Aroostook | 693 | 68,824 | 1.01% | 27.9 | 33.3 | 27.6 | 38.7 | 2 | State |
| Cumberland | 3,000 | 289,361 | 1.04% | 28.8 | 32.9 | 35.1 | 25.8 | 2 | Local |
| Franklin | 115 | 30,072 | 0.38% | 35.7 | 32.2 | 37.4 | 22.6 | 2 | State |
| Hancock | 417 | 54,659 | 0.76% | 35.7 | 33.6 | 55.6 | 20.4 | 2 | State |
| Kennebec | 932 | 119,980 | 0.78% | 35.6 | 32.6 | 55.9 | 24.7 | 1 | Local |
| Knox | 430 | 39,855 | 1.08% | 31.6 | 32.6 | 57.0 | 17.9 | 2 | State |
| Lincoln | 315 | 33,969 | 0.93% | 42.5 | 34.1 | 46.0 | 11.7 | 2 | State |
| Oxford | 323 | 57,202 | 0.56% | 33.7 | 33.3 | 48.9 | 21.7 | 2 | State |
| Penobscot | 1,400 | 152,692 | 0.92% | 29.5 | 33.4 | 44.7 | 29.6 | 2 | Local |
| Piscataquis | 39 | 16,935 | 0.23% | 35.9 | 31.0 | 51.3 | 20.5 | 2 | Local |
| Sagadahoc | 210 | 35,113 | 0.60% | 28.1 | 32.0 | 42.4 | 17.6 | 1 | State |
| Somerset | 285 | 50,745 | 0.56% | 25.6 | 36.4 | 51.2 | 27.0 | 2 | Local |
| Waldo | 352 | 39,155 | 0.90% | 32.4 | 34.1 | 56.8 | 11.7 | 2 | State |
| Washington | 313 | 31,625 | 0.99% | 41.9 | 35.8 | 47.3 | 18.9 | 2 | Local |
| York | 1,038 | 201,169 | 0.52% | 32.5 | 32.1 | 54.0 | 22.5 | 1 | State |

**Figure 1.**
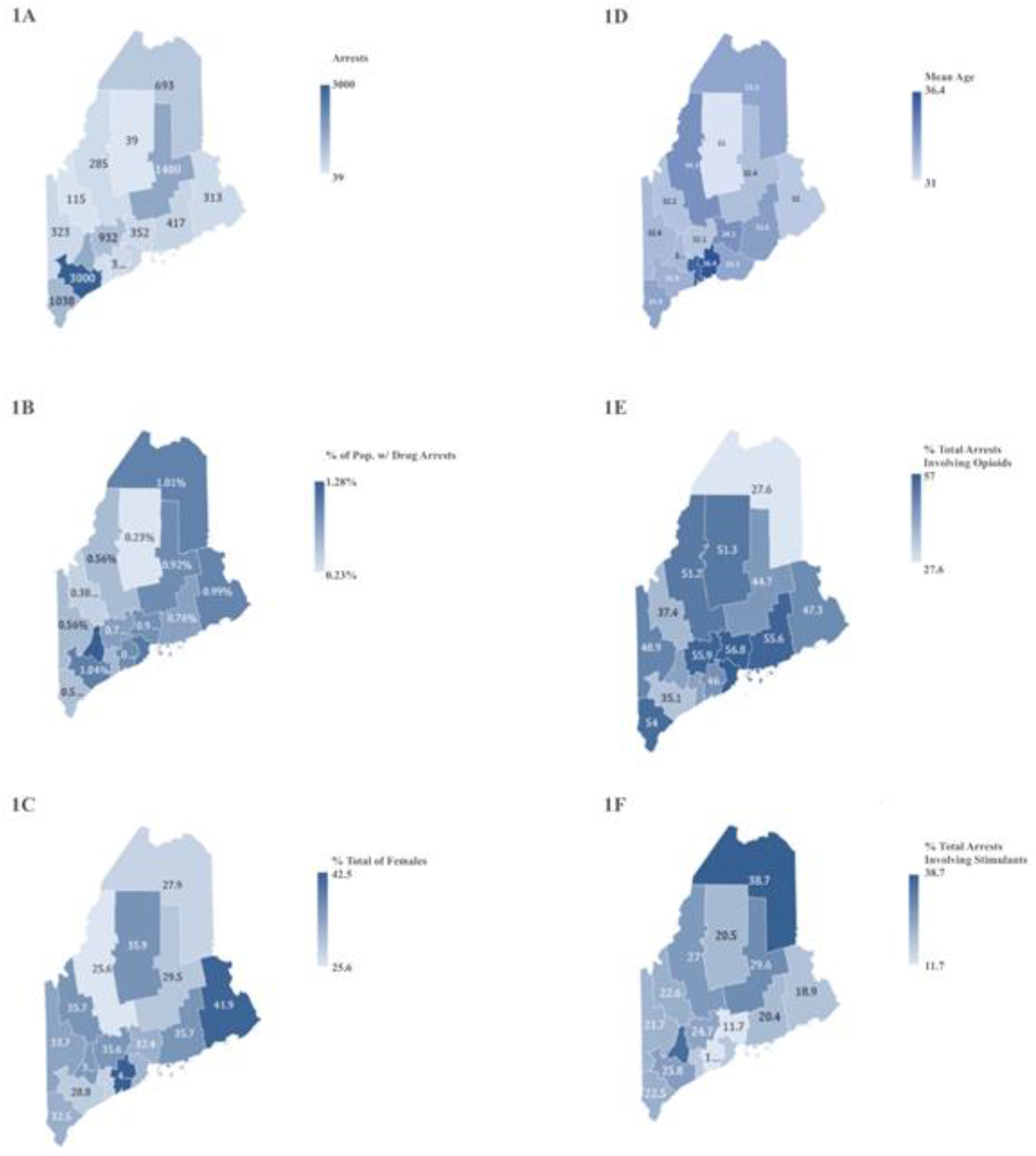
Maine Division Alert Program heat maps show total number of arrests for illicit and prescription drugs (A), percent of the population arrested for illicit pharmaceutical drug (B), percent of females arrested for illicit pharmaceutical drug use (C), mean age of females arrested for illicit pharmaceutical drug use (D), percent of total arrests involving illicit opioid use (E), and total arrests involving illicit stimulant use (F) corrected for population in 2015.

Opioid-related arrests should be considered in the context of trends in the legal distribution of opioids for medical use (pain & OUD) in Maine. Medical distribution of opioids, as defined by MME, increased in 2008 (2,944.8 kg), peaked in 2010 (3,207.5 kg) and declined through 2017 (2,236.0 kg). There was an accelerated decline from 2016 - 2017. Opioids primarily used for pain accounted for one-third of the total MME distributed in Maine in 2008 (32.4%). Distribution of analgesic opioids increased to 36.5% in 2012 before receding to 26.2% in 2017 (Figure 2).

**Figure 2.**
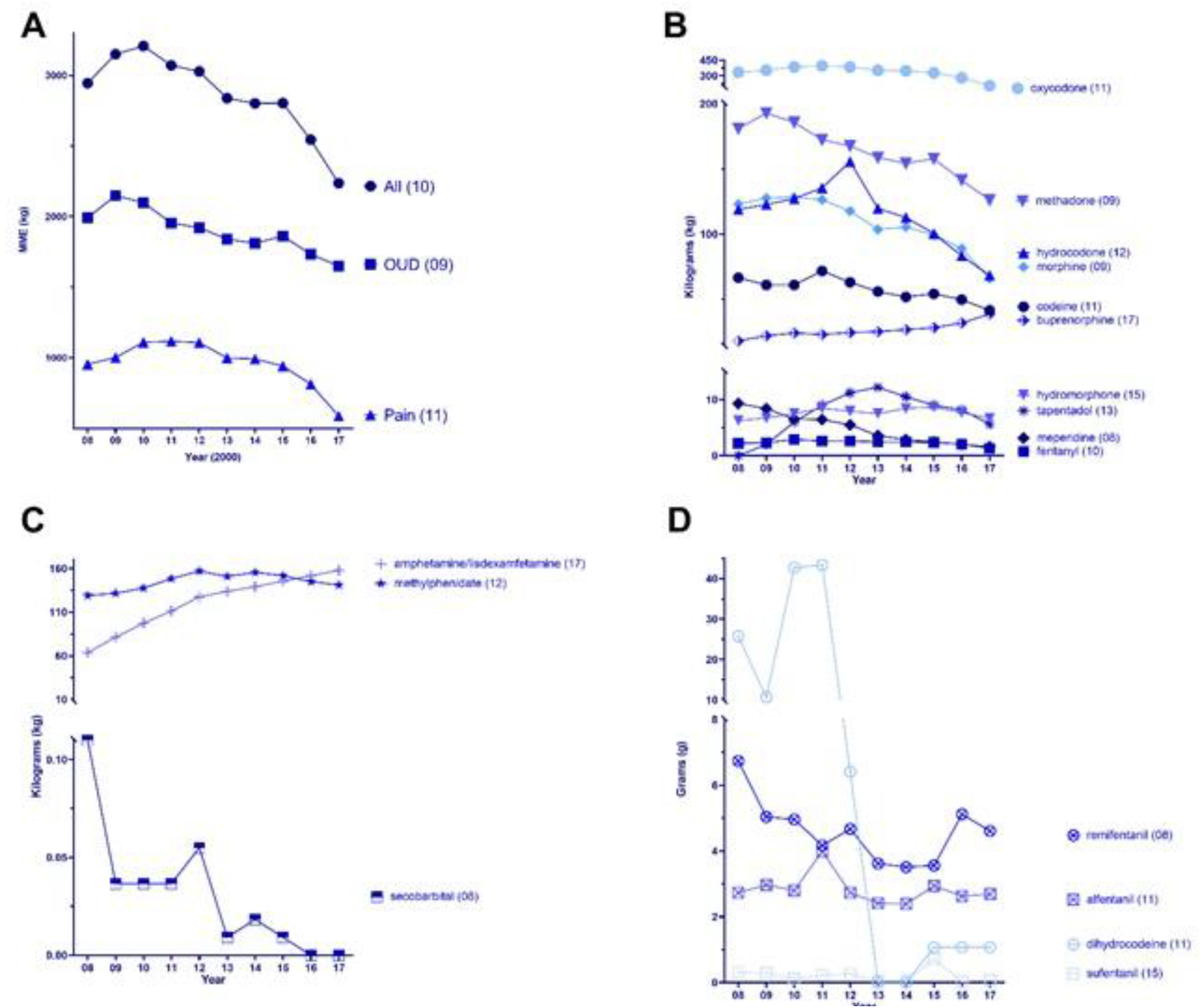
Opioids for pain (oxycodone, fentanyl, morphine, hydrocodone, hydromorphone, oxymorphone, tapentadol, codeine, meperidine), Opioid Use Disorder (OUD, methadone and buprenorphine), and all eleven agents in their morphine milligram equivalents in kilograms per year totaled (A). Raw weights of the most common opioids (B), stimulants (C), and barbiturates (D) in kilograms as reported by the Drug Enforcement Administration in Maine from 2008 to 2017. Percent change relative to the peak year is shown in parentheses.

Figure 2B shows the dynamic changes in distribution of individual opioids standardized to MME. With the exception of buprenorphine distribution (which doubled since 2008), all other prescription opioids have been decreasing. Relative to their peak years over the past decade, hydromorphone, methadone, codeine, oxycodone, and morphine have undergone moderate distribution reductions. Oxymorphone, tapentadol, fentanyl, hydrocodone, and meperidine had large reductions. Lisdexamfetamine distribution increased 6.9-fold since 2008 (9.8 kg). Methylphenidate distribution had a small (−10.1%) decline since peaking in 2012 (157.0 kg). Amphetamine distribution exhibited a large increase (+67.9%) since 2008 (54.0 to 90.7 kg). Pentobarbital distribution remained relatively constant over the decade (79.9 kg in 2008 and 80.9 kg in 2017) while secobarbital distribution precipitously decreased from rare (109.9 g in 2008) to unavailable (0.0 g in 2016 and 2017, not shown).

Further analysis was completed on eleven opioids, expressed as a percent of the total MME, in 2017 by 3-digit zip code which revealed pronounced variations (Figure 3). Opioids used primarily for OUD accounted for the vast majority of opioids distributed in the Bar Harbor (75.0%), Rockland (85.2%), and Bangor (87.4%) zip codes, likely due to the presence of one or more methadone treatment programs in Rockland and Bangor. Methadone was responsible for one-fifth or less of opioids in York (21.0%), Boothbay Harbor (12.6%), and Presque Isle (8.6%, Figure 3B). Hydrocodone accounted for an eightfold greater portion of the total in Presque Isle (9.9%) relative to Portland (1.1%).

**Figure 3.**
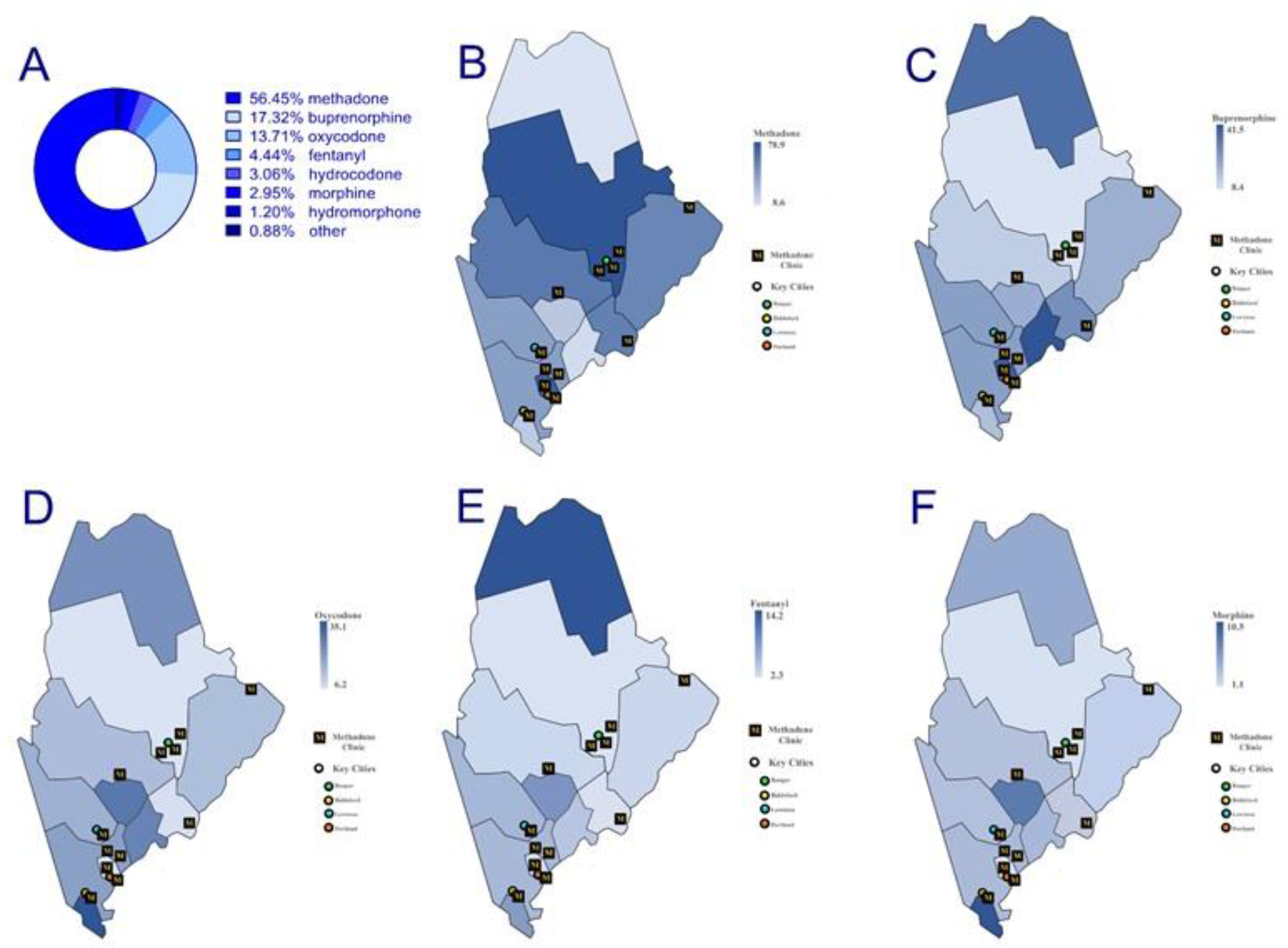
Percent of the total morphine mg equivalent statewide (A) and by 3-digit zip codes (B-F) in 2017 as reported to the Drug Enforcement Administration’s Automation of Reports and Consolidated Ordering System. Other: tapentadol, meperidine, oxymorphone and codeine.

## DISCUSSION

The first two objectives of this study were to examine county and demographic differences in controlled substance misuse in Maine. The DAP provided information which complemented Prescription Monitoring Programs (PMP) for patient care. However, the DAP was also invaluable for research and could complement other data sources like self-report from the National Household Survey on Drug Use and Health or Monitoring the Future, emergency room reports, and drug seizures.^12^ A key theme of prior reports was the substantial diversity of substances implicated in arrests beyond the “usual suspects” of heroin, oxycodone, hydrocodone, illicit fentanyl, methamphetamine, and cocaine to also include hundreds of arrests for the benzodiazepines alprazolam and clonazepam, marijuana, and miscellaneous non-controlled prescription pharmaceuticals like gabapentin and quetiapine.^9^

There were almost ten-fold more arrests from 2014 to 2017 involving buprenorphine (812) than methadone (82). The disparity was striking given that, on an MME basis, methadone was distributed state-wide over three-fold more than buprenorphine. However, they can be at least partially explained by considering the distribution of methadone for patients with OUD. Typically, addiction medicine providers dispense onsite at methadone clinics. To date, there are twelve methadone clinics spanning nine counties in Maine.^13^ These clinics contain providers that specialized in the medication treatment of patients with opioid use disorder, especially methadone, as well as provide counselling services.

A high rate of buprenorphine arrests may still be surprising, given its pharmacology as a partial mu receptor agonist. Although buprenorphine has a considerably more favorable safety profile than methadone (LD50 = 235 vs 23 mg/kg iv in rats)^14^, there were still eleven-thousand poison control reports involving buprenorphine, primarily as a monotherapy product, among children and adolescents (< 19) from 2007 to 2016.^15^ Buprenorphine was identified in 22 drug deaths in Maine in 2017.^16^ In other states, such as Wisconsin, law enforcement have identified an increased number of cases of driving under the influence of buprenorphine, often with benzodiazepines.^17^ Although both methadone and buprenorphine are considered efficacious for OUD treatment, the provision of methadone for OUD is considerably more restricted than buprenorphine. A systematic review identified better retention in OUD treatment with methadone than buprenorphine/naloxone.^18^ Methadone accounted for one-eighth of the total opioid MME in Presque Isle and Boothbay Harbor but three-quarters in Bangor and Portland. Maine methadone treatment programs increased from nine in 2008 to ten in 2017.^10^ However, their distribution is not regionally uniform with four in Cumberland County and three in Penobscot.^13^ This continues to be a barrier to providers. Almost two-thirds of opioid treatment program (OTP) patients resided in just three counties (Penobscot = 33%, Cumberland = 22%, Washington = 8%).^19^ Although there is room for improvement, Maine did rank second in the US for the most waivered physicians for buprenorphine.^20^

The pronounced reductions in opioid distribution over the past decade has been reported previously.^21^ Perhaps what is more novel is the rate of change in opioid use, particularly from 2015 to 2017. The liberalization of opioid prescribing practices over the 2000s was the unfortunate result of a confluence of factors working together.^7^ We suspect that the return to more judicious and evidence-based use of opioids, particularly minimizing use in patients with chronic non-cancer pain, may also reflect the convergence of multiple policy and incentive changes at the federal, state, and local level. Maine’s “An Act to Prevent Opiate Abuse by Strengthening the Controlled Substances Prescription Monitoring Program,” was unusual for a state prescribing law in that it included fiscal penalties for non-adherence. Maine had a significantly greater decrease in prescription opioids than several other New England and mid-Atlantic states (CT, MA, NY, PA, RI, VT) that enacted similar opioid prescribing laws but that lacked penalties.^21^ Drug related deaths involving prescription opioids in Maine declined by 37% from 2017 to 2018.^16^

Three other findings are noteworthy. The substantial elevations in lisdexamfetamine and other amphetamine distribution extend upon earlier national data.^7^ ARCOS, and DAP, do not provide information about whether the original source of these stimulants was for ADHD in children or adults, obesity, post traumatic brain injury, or another indication. The ratio of males to females for drug related deaths in Maine was 2.51 to 1 in 2017^5^ and 2.45 to 1 in 2018.^16^ Among arrestees, the ratio was similar (2.14 to 1). Together with other arrest data, these findings indicated that sex differences in drug misuse may gradually be eroding.^17^

Some strengths and limitations of these datasets are noteworthy. The DAP was unique to Maine and, to our knowledge, has not been emulated elsewhere. Reporting to the DAP was voluntary and some agencies (e.g. tribal police) very infrequently submitted arrest information which is a caveat in interpreting Figure 1. It is important to emphasize that there are key differences between an arrest and a subsequent conviction and data with regard to conviction is unknown. It is also unknown how often field tests whose specificity is questionable were employed to determine the presumptive substance identity.^22^ Finally, there is a potential for bias in DAP with regard to populations who are more likely to be surveilled by law enforcement and subsequently arrested, such as those with a history of opioid overdose or diagnosed OUD. There are also socioeconomic and racial disparities that might be contributing to bias. Unlike PMPs, ARCOS is comprehensive in its coverage of Schedule II substances. ARCOS reports by substance weight instead of using perhaps more intuitive units of analysis like prescriptions. Methadone is the most prevalent opioid by MMEs.^3,23^ The arguably well intentioned 42 CFR Part 2 which prevented methadone when used by a methadone treatment programs from being entered into the state PMP results in a key caveat when interpreting pharmacoepidemiological research.^8,24^ Some have advocated that this federal regulation be updated.^25^

In conclusion, the DAP was unique to Maine and was an important harm-reduction tool, but it was not without limitations. We are cautiously optimistic that the DAP might be worthy of consideration in other states in order to improve information transfer from law enforcement to health care providers.

## Data Availability

Arrest data is sensitive. Our data use agreement precludes sharing Diversion Alert data. The Drug Enforcement Administration's ARCOS data is publically available online.

https://www.deadiversion.usdoj.gov/arcos/retail_drug_summary/index.html

## Acknowledgements

This research was supported by the Fah-Beck Fund for Research and Experimentation and the Health Resource and Services Administration (D34HP31025). Maureen J. Murtha and Salomey O. Mensah facilitated the summer team.

## Disclosures

BJP has osteoarthritis research supported by Pfizer. A member of his household was previously employed by Garlic Acres, a company that produced medical cannabis products. SDN has consulted for Shire. MF and CED were employed by the Diversion Alert Program. The other authors have no disclosures.

**Word Count:** Abstract: 250/250; Manuscript: 2,478/3,000; Figures: 3; Tables: 1, Sup Fig: 1

---

**Supplemental Figure 1.**
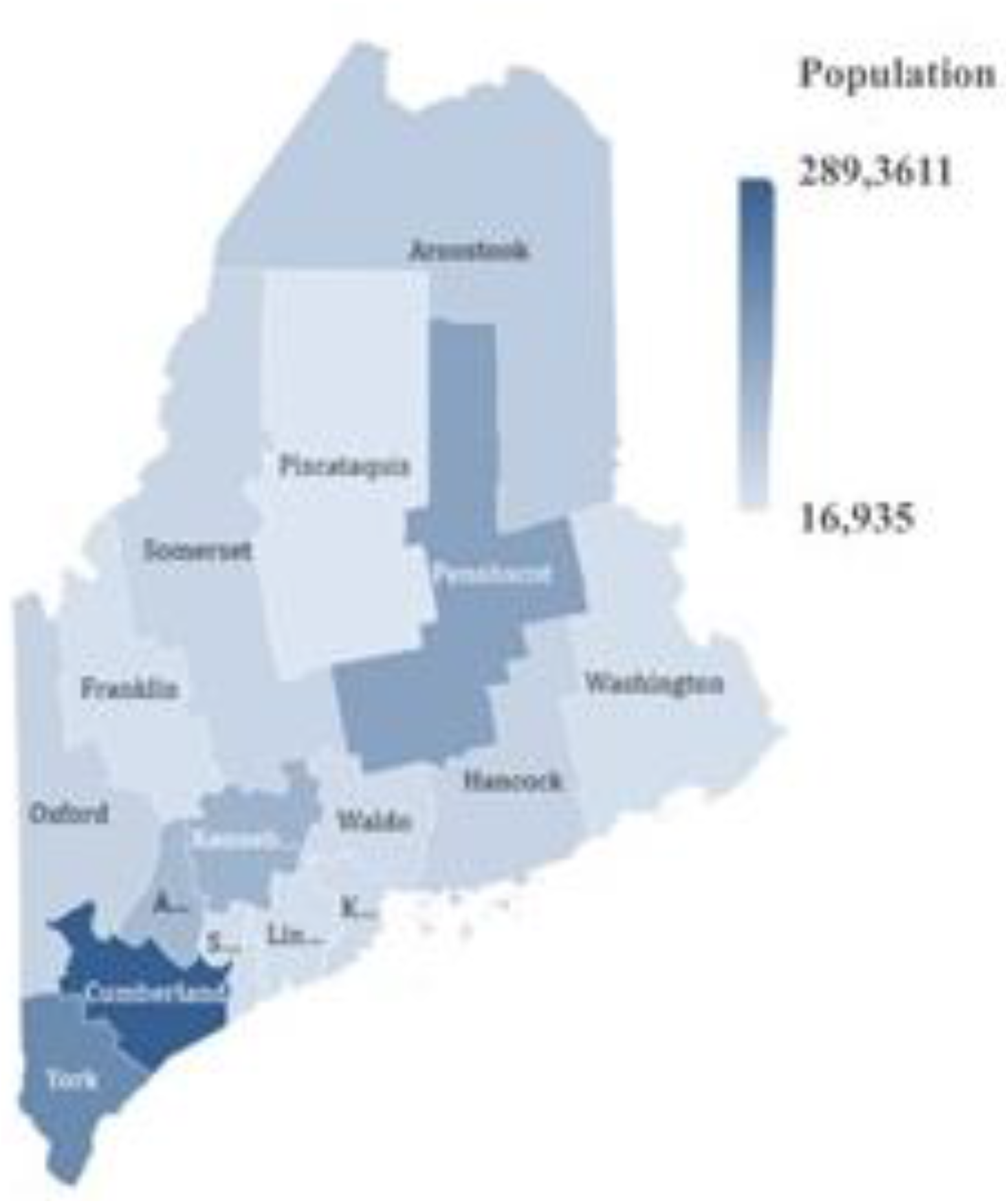
Heat map of Maine’s population size by county in 2015.

